# Rapid Detection of Novel Coronavirus (COVID-19) by Reverse Transcription-Loop-Mediated Isothermal Amplification

**DOI:** 10.1101/2020.02.19.20025155

**Authors:** Laura E. Lamb, Sarah N. Bartolone, Elijah Ward, Michael B. Chancellor

## Abstract

Novel Corona virus (COVID-19 or 2019-nCoV) is an emerging global health concern that requires a rapid diagnostic test. Quantitative reverse transcription PCR (qRT-PCR) is currently the standard for COVID-19 detection; however, Reverse Transcription Loop-Mediated Isothermal Amplification (RT-LAMP) may allow for faster and cheaper field based testing at point-of-risk. The objective of this study was to develop a rapid screening diagnostic test that could be completed in under 30 minutes. Simulated patient samples were generated by spiking serum, urine, saliva, oropharyngeal swabs, and nasopharyngeal swabs with a portion of the COVID-19 nucleic sequence. The samples were tested using RT-LAMP as well as by conventional qRT-PCR. Specificity of the RT-LAMP was evaluated by also testing against other related coronaviruses. RT-LAMP specifically detected COVID-19 in simulated patient samples. This test was performed in under 30 minutes. This approach could be used for monitoring of exposed individuals or potentially aid with screening efforts in the field and potential ports of entry.

## Introduction

The recent outbreak of Novel Coronavirus (COVID-19) has generated global concern given its rapid spread in multiple countries and possible fatal progression of the infection. Initially, many patients reported exposure at a large seafood and animal market in Wuhan, China, suggesting animal-to-person transmission of the virus. However, since then many patients have reported no exposure to animal markets, indicating that person-to-person transmission is occurring. There is currently no vaccine or targeted therapeutic for COVID-19.

COVID-19 is difficult to diagnose early in infection as patients can remain asymptomatic or present with non-specific flu-like clinical symptoms including fever, cough, or shortness of breath. Symptoms may appear in as few as 2 days or up to 2 weeks after exposure.^1^ Quantitative reverse transcription PCR (qRT-PCR) for COVID-19 in serum or respiratory samples is currently the standard for diagnostic molecular testing. However, this requires expensive equipment and trained personnel.

Reverse transcription loop-mediated isothermal amplification (RT-LAMP) is a one-step nucleic acid amplification method based on PCR technology that has been used to diagnose infectious diseases.^2^ RT-LAMP has several advantages including that it has high specificity and sensitivity, can be done in less than an hour, can work at various pH and temperature ranges which is advantageous for clinical samples,^3^ and that the reagents are relatively low cost and can be stable at room temperature. We have previously used this method to detect zika virus in clinical serum and urine samples as well as mosquitos.^4, 5^

This study describes a RT-LAMP methodology that can detect COVID-19 in simulated patient samples in under 30 minutes. The test could be used at the point-of-care by field and local personnel for the rapid diagnosis of individuals for optimal treatment, isolation, and rapid contact tracking as well as the investigation of outbreaks of unknown respiratory diseases.

## Methods

### RT-LAMP primer design

The consensus sequences of 23 different COVID-19 strains (EPI_ISL_403929, EPI_ISL_403930, EPI_ISL_403931, EPI_ISL_402129, EPI_ISL_402130, EPI_ISL_402132, EPI_ISL_402128, EPI_ISL_402124, EPI_ISL_402127, EPI_ISL_402121, EPI_ISL_402119, EPI_ISL_402123, EPI_ISL_403962, EPI_ISL_403963, EPI_ISL_404227, EPI_ISL_404228, EPI_ISL_403932, EPI_ISL_403933, EPI_ISL_403934, EPI_ISL_403935, EPI_ISL_403937, EPI_ISL_403936, EPI_ISL_406596, EPI_ISL_406844, EPI_ISL_407079, EPI-ISL_407214) were established when aligned with Lasergene MegAlign (DNASTAR) to identify areas of sequence conservation. To improve specificity, areas of divergence of COVID-19 (GenBank MN908947) with the sequence-related coronavirus, Bat Severe Acute Respiratory Syndrome (SARS)-like coronavirus (GenBank KY417152.1), were identified using BLAST 2 (Basic Local Alignment Search Tool; NCBI). An area of GenBank MN908947 sequence that had high homology with other COVID-19 strains and was divergent for Bat SARS-like coronavirus was then targeted for RT-LAMP primer design. RT-LAMP primers were designed using LAMP Designer 1.15 (Premier Biosoft), and blasted using Primer-BLAST (NCBI) against genomes of interest. Primer selection was prioritized as described in “A Guide to LAMP primer designing” (https://primerexplorer.jp/e/v4_manual/). In addition, primers were selected to not have four guanines in a row to prevent the formation of tetraplex structures which could disrupt the RT-LAMP reaction. RT-LAMP primers are listed in Table 1. Primer sets include an outer forward primer (F3), outer backward primer (B3), forward inner primer (FIP), backward inner primer (BIP), loop forward primer (LF), and loop backward primer (LB). Primers were ordered from Integrated DNA Technologies and the FIPs and BIPs are HPLC purified.

**Table 1.**
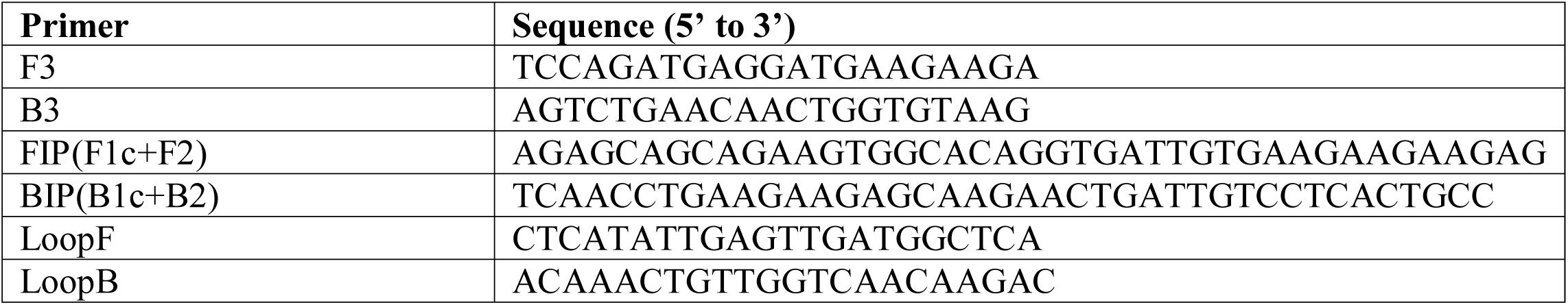
RT-LAMP primers for COVID-19 detection.

### Control strain details

COVID-19 PCR-standard was designed from nucleotide 2941-3420 of the COVID-19 Wuhan-Hu-1 complete genome (MN908947). The COVID-19 ssDNA control fragment was synthesized through Invitrogen GeneArt. An oligo of GenBank MN908947.3 was used as there were no commercial sources for a COVID-19 control plasmid at the time this paper was prepared. The fragment was reconstituted to 10ng/µL and used for LAMP and qPCR experiments. RT-LAMP and PCR controls for Middle East Respiratory Syndrome (MERS), Betacoronavirus England-1 (BtCoV), and Murine hepatitis virus (MHV) were similarity made. We were unable to make a RT-LAMP and PCR control for any SARS sequences as this is controlled under European Union export regulations.

### Patient samples

All studies were approved by Beaumont Health’s Institutional Review Board (IRB approval # 2020-040 and 2016-044). All study participants gave their informed consent to participate. All experiments were performed in accordance with the ethical standards noted in the 1964 Declaration of Helsinki and its later amendments. Samples from multiple healthy, consenting, adult volunteers were tested from either fresh or frozen state for each assay. Fresh samples were kept on ice until analysis. For whole mouth saliva and oropharyngeal swabs, participants refrained from eating or drinking anything other than water for 5 minutes prior to collection and rinsed their mouths with water prior to collection. Oropharyngeal swab and nasopharyngeal swabs samples were collected using a Flocked sterile plastic swab applicator which was placed in a Universal Viral Transport for Viruses, Chlamydiae, Mycoplasmas, and Ureaplasmas vial (Bector, Dickinson and Company). To demonstrate our point-of-care RT-LAMP assay and to test possible interference with agents in serum, urine, saliva, oropharyngeal swab, and nasopharyngeal swab collection, various concentrations of COVID-19 oligo were spiked in samples for test validation. Water was used as a no template control.

### COVID-19 RT-LAMP Primer Specificity

Percent mismatch for RT-LAMP primers was determined by aligning the COVID-19 genome (Wuhan City, Hubei, 2019-12-26, GenBank ID MN908947) to 22 other sequenced COVID-19 strains from various locations around the world using the Global Initiative on Sharing All Influenza data (GISAID) EpiFlu database and the BLAST Global Alignment tool.^6, 7^ RT-LAMP primers were also compared to other coronaviruses using the BLAST Global Alignment tool to determine specificity and percent mismatch using the same method. Percent mismatch was calculated using the following equation:

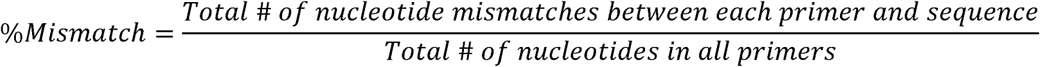

### RT-LAMP

All set-up and execution of RT-LAMP reactions were done in an enclosed room using designated pipettes and filter tips. Analysis and imaging took place in separate rooms to prevent contamination. RT-LAMP reactions were executed in a total volume of 25μL of 1x isothermic amplification buffer, 1·4 mM dNTPs, 8 mM MgSO_4_, 1·6 μM FIP/BIP, 0·2 μM F3/B3, 0·4 μM FL/BL primers, 0·32 U/μL *Bst 2*·*0*, 1 U/μL Antarctic Thermolabile UDG, and 0·6 U/μL *WarmStart Reverse Transcriptase* in ddH_2_0. Addition of the uracil-DNA glycosylase (UDG) reduced crossover contamination from previous reactions. Reactions were set-up on ice, incubated at 63°C for 30 minutes (15-45 minutes have been tested), and then inactivated at 80°C for 10 minutes. In order to optimize visualization of positive reactions, 2 μL SYBR Green I was added to reactions at a 1:10 dilution. All experiments were replicated 3-5 times.

### qRT-PCR

For testing, a 1:5 serial dilution of COVID-19 ssDNA control fragment was made, from 2 ng to 0.204 fg. Nuclease-free ddH_2_O was used as a no template control. No template control or COVID-19 was added to SYBRSelect Master Mix (ThermoFisher), and 10 µM forward and reverse primers with a total reaction volume of 20 µL. Primers used were F 5’-ATTTGGTGCCACTTCTGCTGC −3’ and R 5’-TCACTGCCGTCTTGTTGACCA −3’. Cycling conditions were 50 °C for 2 minutes and 95 °C for 10 minutes, followed by 40 cycles of 95 °C for 15 seconds and 60 °C for 1 minute, performed on a QuantStudio 3 PCR System (Applied Biosystems).

### Analysis of RT-LAMP

1:10 SYBR green I (Life Technologies) dilution was made in TAE buffer, then 2 μL of the SYBR dilution was added to the RT-LAMP reactions. The visual change of color (orange to yellow) was also used to identify positive amplifications. In addition, the SYBR green I PCR tubes were imaged under UV light in the BIO-RAD ChemiDoc XRS+ Imaging System as the reaction creates a fluorescent output. One half of the RT-LAMP reaction was electrophoresed along with Invitrogen Low DNA Mass Ladder on a 2 % agarose gel in 1x TAE buffer (40 mM Tris, 20 mM acetic acid, 1 mM EDTA) at 90 V for 90 minutes. Gels were imaged under UV light using the BIO-RAD ChemiDoc XRS+ Imaging System. Lanes containing a laddered banding pattern were qualified as a positive amplification.

## Results

### RT-LAMP is specific and sensitive for COVID-19

To establish the optimal conditions for RT-LAMP using a COVID-19 PCR-standard designed from nucleotide 2941-3420 of the COVID-19 Wuhan-Hu-1 complete genome (MN908947), several primer sets, ranges of temperatures (57-65°C), and incubation times (15-45 mins) were tested. The best amplification results were obtained at 63 °C for 30 minutes as indicated by a banding pattern after electrophoresis on a gel (Fig 1). Positive reactions containing SYBR Green I could be observed by naked eye by a color change from orange to yellow (Fig 1-top panel), under fluorescent light in response to UV excitation (Fig 1-middle panel), or by laddering pattern of bands after electrophoresis on a gel (Fig 1-bottom panel). The RT-LAMP reaction required all 6 primers to work under the optimized conditions; removing the forward and backward inner primers or the loop primers did not result in a positive result (Fig 1). In order to determine the lower detection limit of the RT-LAMP reaction for COVID-19, a dilution series ranging from 0.204 fg to 10 ng COVID-19 was amplified (Fig 2). The limit of detection was approximately equivalent to 1.02 fg. This dilution series was run in parallel with qRT-PCR using primers that targeted this same region of COVID-19 genome; the qRT-PCR Ct values for the dilution series are reported in Supplemental Table 1.

**Fig 1.**
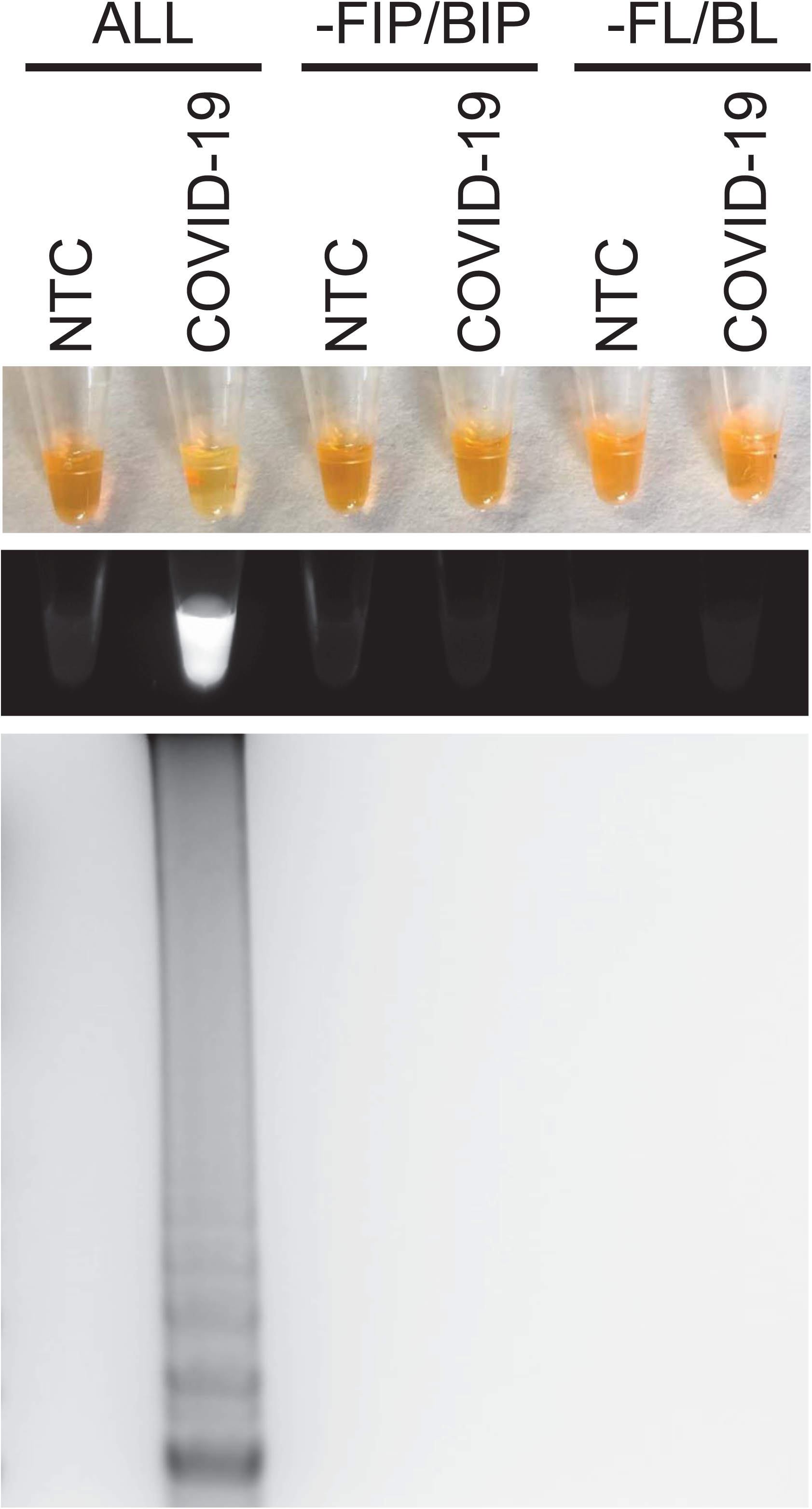
RT-LAMP detection of COVID-19. (A) COVID-19 RT-LAMP amplification of COVID-19 PCR standard (COVID-19; IDT custom oligo) but not no template control (NTC; negative control) as visualized by addition of SYBR Green I (SYBR) by eye (upper panel), fluorescence (middle panel), or gel electrophoresis (bottom panel). All primers (ALL) are required for effective RT-LAMP reaction. Reactions without FIP and BIP (-FIP/BIP) or FL and BL (-FL/BL) primers resulted in a negative RT-LAMP reaction.

**Fig 2.**
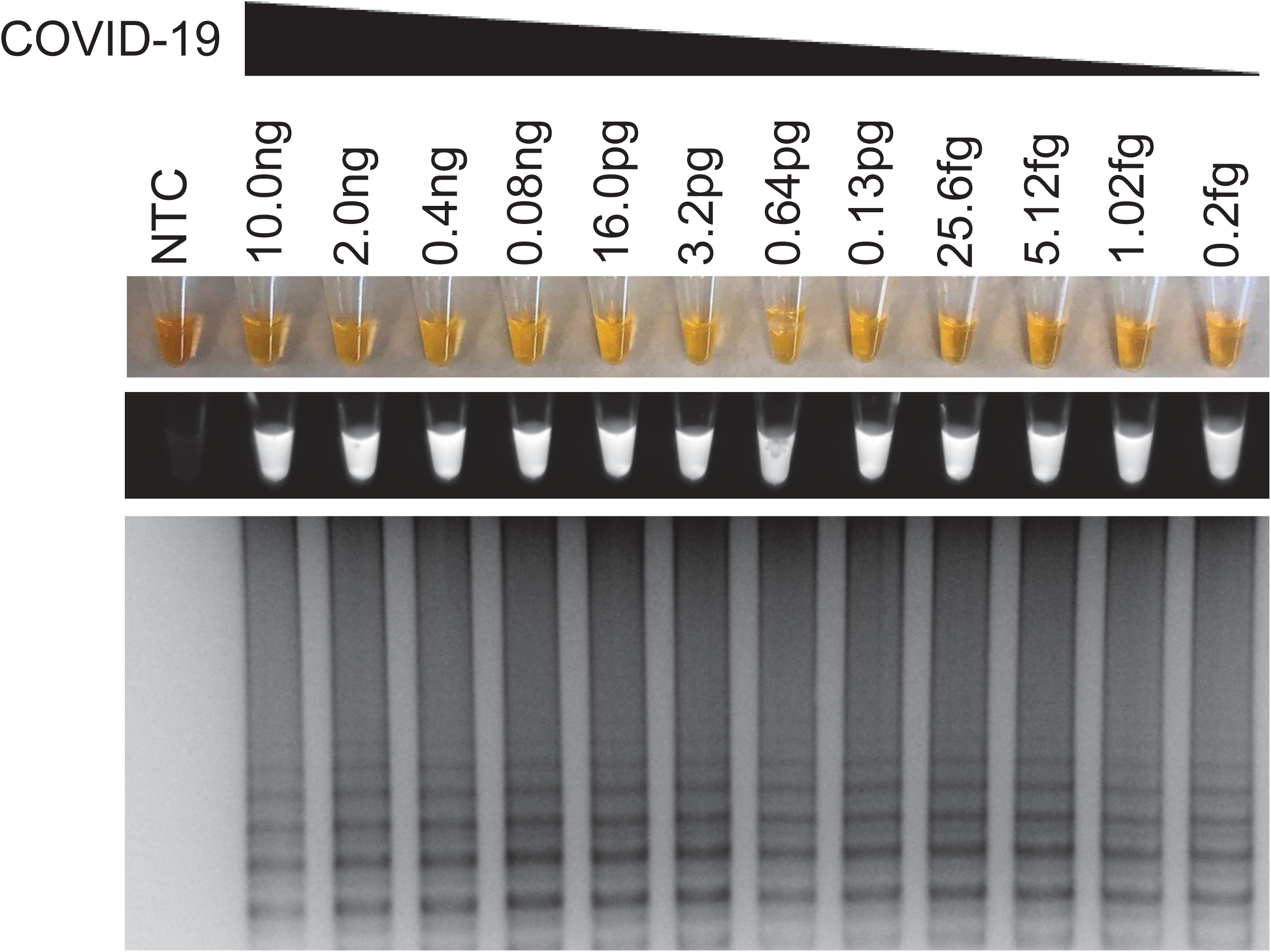
COVID-19 RT-LAMP sensitivity for COVID-19. Sensitivity assessment of COVID-19 RT-LAMP using serial dilutions of COVID-19 PCR Standard from 10·0 ng/reaction to 0·2 fg/reaction as visualized by addition of SYBR Green I by eye (upper panel), fluorescence (middle panel), or gel electrophoresis (bottom panel). NTC: No template control (negative control).

### COVID-19 Detection in Simulated Clinical Samples with RT-LAMP

To demonstrate the clinical utility of this system for COVID-19 detection, we spiked various human specimens with COVID-19, Middle East Respiratory Syndrome (MERS), Betacoronavirus England-1 (BtCoV), or Murine hepatitis virus (MHV). MERS, BtCoV and MHV spiked samples were used to test the specificity of the RT-LAMP assay. Spiked specimens included serum, urine, saliva, oropharyngeal swabs, and nasopharyngeal swabs as we wanted to verify that substances present in these samples would not interfere with RT-LAMP. Samples were directly used for RT-LAMP without performing nucleic acid isolation. Only samples containing COVID-19, but not MERS, BtCoV, or MHV, had positive RT-LAMP reactions indicating specificity of the reaction for COVID-19 (Fig 3). Furthermore, RT-LAMP could be successfully completed using human serum, urine, saliva, oropharyngeal swabs, and nasopharyngeal swabs.

**Fig 3.**
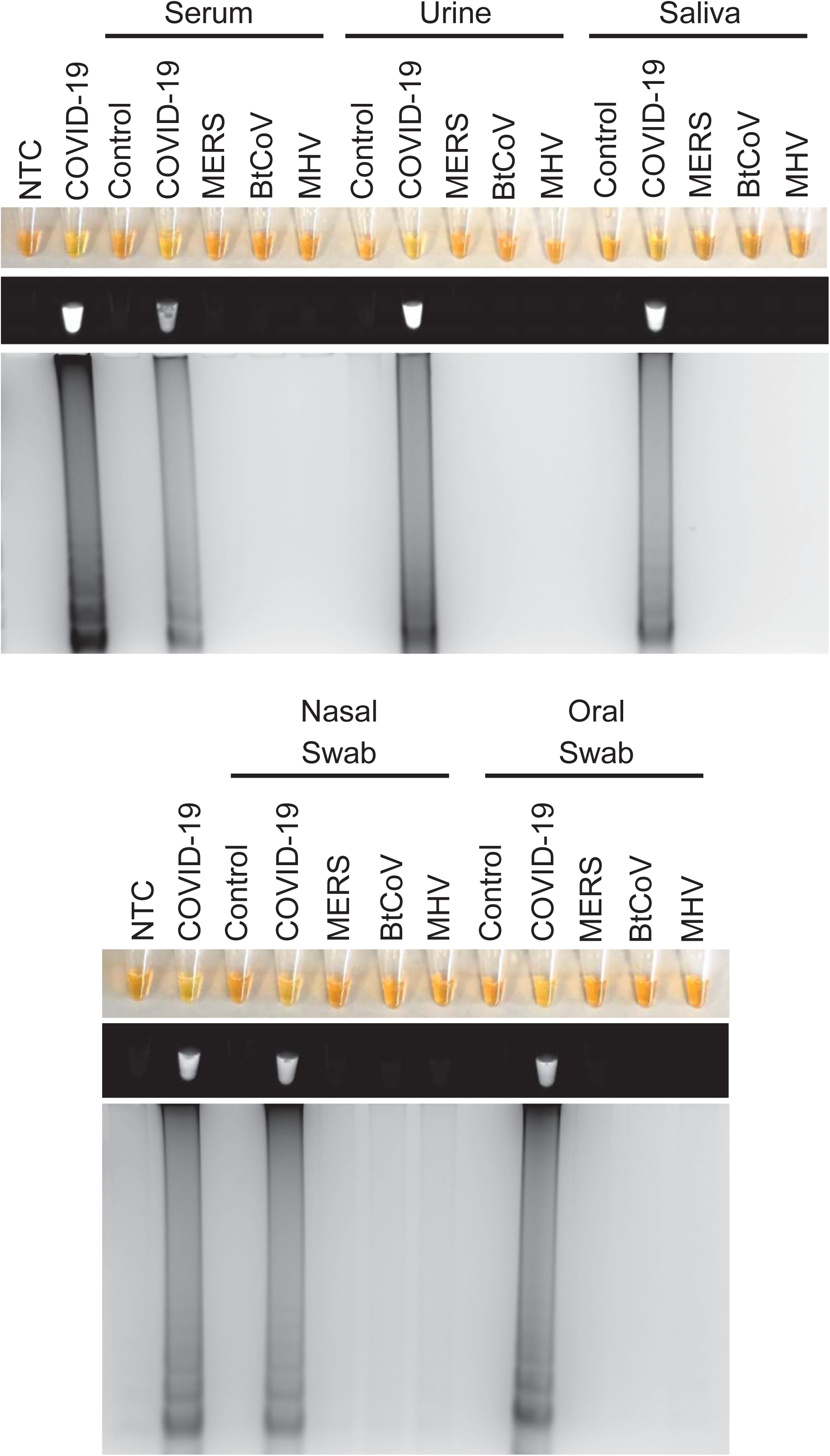
COVID-19 RT-LAMP specificity for COVID-19 in simulated patient samples. Specificity assessment of COVID-19 RT-LAMP in control samples (control) or samples spiked with COVID-19, MERS, BtCoV, MHV PCR standards (IDT custom oligos) as visualized by the addition of SYBR Green I by eye (upper panel), fluorescence (middle panel), or gel electrophoresis (bottom panel). Types of human samples tested included serum, urine, saliva, nasopharyngeal swabs (nasal swab), or oropharyngeal swabs (oral swab). NTC: No template control (negative control). 3-5 different patient samples were tested for each condition with one representative patient being shown.

### RT-LAMP is Specific for COVID-19

The sequence of the RT-LAMP primers were also compared to aligned sequences of various strains of Severe Acute Respiratory Syndrome coronaviruses (SARS) or coronaviruses commonly associated with the common cold (human coronavirus strains 229E, NL63, HKU1, or OC43). These other coronaviruses had 27-54% nucleotide mismatch with our RT-LAMP primers, making it highly unlikely they would give a positive RT-LAMP result, supporting the specificity of this assay for COVID-19 (Table 2). Given that viruses are prone to genetic mutation, we likewise examined if RT-LAMP primers had any mismatch with 27 different isolated strains of COVID-19 from various locations. There was 0% mismatch with all the strains examined, suggesting that these RT-LAMP primers would identify all 27 strains of COVID-19 examined (Supplemental Table 2).

**Table 2.** RT-LAMP primers alignment with other coronaviruses.

| **Virus** | **GenBank ID** | **% Mismatch** |
| --- | --- | --- |
| COVID-19 | MN908947 | 0 |
| Bat SARS-like CoV 2015 | MG772933.1 | 27.22 |
| Bat SARS-like CoV 2017 | MG772934.1 | 30.77 |
| Bat SARS CoV RM1/2004 | KY417144.1 | 40.83 |
| SARS CoV ZS-C | AY395003.1 | 42.60 |
| Civet SARS CoV SZ16/2003 | AY304488.1 | 39.64 |
| SARS CoV | NC_004718.3 | 40.24 |
| SARS CoV MA15 | FJ882957.1 | 40.24 |
| Middle East Respiratory CoV | NC_019843.3 | 52.07 |
| Belacoronavirus England 1 | NC_038294.1 | 54.44 |
| Murine hepatitis virus | NC_001846.1 | 53.25 |
| Human Coronavirus 229E | NC_002645.1 | 51.48 |
| Human Coronavirus NL63 | NC_005831.2 | 54.44 |
| Human Coronavirus HKU1 | NC_006577.2 | 53.25 |
| Human Coronavirus OC43 | NC_006213.1 | 49.11 |

## Discussion

Given the rapid emergence of COVID-19 and the severe complications that can result including acute respiratory distress syndrome (ARDS) and pneumonia, improved diagnostic options that are fast, reliable, easy, and affordable are required. This is critical since COVID-19 infection can rapidly progress from hospital admission to ARDS in as few as 2 days, and COVID-19 infection can be fatal.^1^ Conventional qRT-PCR, while specific and sensitive, must be done by trained personnel on specialized equipment at a qualified laboratory. Since this disease is spreading rapidly, centralized labs may have trouble keeping up with testing demands or may need an alternative approach if qRT-PCR kits are not available. This feasibility study demonstrated that RT-LAMP allows rapid detection of COVID-19 in a variety of common human specimens collected for clinical testing, including serum, urine, saliva, oropharyngeal swabs, and nasopharyngeal swabs.

Currently, clinical testing for COVID-19 is done by central testing laboratories, which may take one or more days. This study sought to improve upon this by developing a potential point-of-care test. Point-of-care testing has several advantages for emerging infectious diseases like COVID-19 and must be easy to use, inexpensive, fast, and require little if any laboratory infrastructure while maintaining sensitivity and specificity. RT-LAMP meets these requirements and therefore has large value for screening and testing for COVID-19 in potentially exposed populations.

We sought to determine if this RT-LAMP assay worked in a range of different samples that might be collected in a clinical setting or as a possible non-invasive screening tool. This is important since it may not be feasible to collect serum from all patients, especially patients who are critically sick, dehydrated, the elderly, children, and neonates. Personnel trained in collecting blood specimens may also not be available, Furthermore, biological samples may contain chemicals that can inhibit nucleic acid assays if the sample is tested directly without first isolating the RNA. RT-LAMP worked in all the human sample types tested, and in samples from several individuals. We previously demonstrated that RT-LAMP for ZIKV does not require prior RNA isolation from the samples.^4, 5^ Thus, we used unprocessed urine or serum samples in this study, which saves considerable time and reduced costs. In this RT-LAMP assay for COVID-19, the urine samples gave stronger RT-LAMP signals than the serum samples when spiked with identical amounts of COVID-19; however the amount of virus present in an infected individual will likely vary between different biological specimens and over the time course of the infection. Furthermore, we observed faint background banding by gel electrophoresis in oropharyngeal swabs, suggesting there may be some factor in this specimen that results in this. However, due to the specificity of the primers, we believe this does not interfere with correct interpretation of the RT-LAMP reaction, especially since the COVID-19 spiked sample can be correctly identified through color change and fluorescence.

Primers were designed for a conserved span of COVID-19 sequence that was found in 22 isolated COVID-19 strains but was a sequence that was also divergent from related coronavirus SARS. Given that all the isolated strains of COVID-19 thus far have shown very little genetic differences, we anticipate that this RT-LAMP will detect COVID-19 with the same level of confidence as qRT-PCR. This COVID-19 RT-LAMP assay was highly specific as it did not give a positive result for MERS, BtCoV, and MHV and had significant mismatch with numerous strains of SARS and common cold associated human coronoavirus strains 229E, NL63, HKU1, or OC43. We tested a range of temperatures from 57°C to 65°C; all the temperatures gave comparable results, indicating that this RT-LAMP reaction has a large temperature range with an optimal temperate of 63 °C. We also tested a range of incubation times from 15-45 minutes. The optimal time for detection of RT-LAMP products was 30 minutes, however COVID-19 RT-LAMP products were detectable by UV light excitation or banding patterns on gels in as little as 15 minutes.

This study has several limitations. First, COVID-19 is Biosafety level 3 so our laboratory was unable to work directly with the virus or with infected samples. As such, all the experiments presented here used a nucleotide oligo of COVID-19 corresponding to the GenBank MN908947 sequence. Similarly, we were unable to directly test related coronaviruses and instead used nucleotide oligos from the same region of those viruses. However, these do represent a proof-of-feasibility for this assay, and the primers were further evaluated for specificity by BLASTing them to related coronavirus sequences. Although RT-LAMP reactions are highly specific, it is not a quantitative test. However other groups are working at improving the read outs of RT-LAMP assays including the use of smartphone-integrated sensors to make interpretation of the assay even more user-friendly.^8^ RT-LAMP reactions can have a higher rate of false positives compared to qRT-PCR; we did not experience this in any of our no template negative control reactions, negative control samples, or other samples containing other coronaviruses. We also took the precautions of having a lateral work flow for experiments and we included a Thermolabile Uracil-DNA Glycosylase (UDG) in all reactions to prevent possible carry-over contamination from previous reactions. This study was not powered to determine sensitivity in a clinical population. Lastly, this is a rapidly developing area of study and all the information presented at the time of publication represent the authors’ best knowledge at the time. As our understanding of COVID-19 continues to develop, this information may change.

## Conclusions

Here we describe a fast and robust assay for detection of COVID-19 in under 30 minutes. This simple assay could be used outside of a central laboratory on various types of biological samples. This assay can be completed by individuals without specialty training or equipment and may provide a new diagnostic strategy for combatting the spread of COVID-19 at the point-of-risk.

## Data Availability

All relevant data are within the paper.

## Acknowledgements

We would like to thank Denise Cunningham and Dr. Girish Nair for their assistance obtaining human samples. We also thank Dr. Bernadette Zwaans for her critical review of manuscript.

## Additional Information

### Author Contributions

LEL and MBC conceived of the work. RT-LAMP primer design was by LEL. Data collection was by SNB and EW. All authors performed data analysis and interpretation. LEL drafted the article. All authors critically revised the article and gave approval of the final version.

### Data Availability

All relevant data are within the paper.

